# COVID-19 among people living with HIV: A systematic review

**DOI:** 10.1101/2020.07.11.20151688

**Authors:** Hossein Mirzaei, Willi McFarland, Mohammad Karamouzian, Hamid Sharifi

## Abstract

This systematic review summarizes the evidence on the earliest patients with COVID-19-HIV co-infection. We searched PubMed, Scopus, Web of Science, Embase, preprint databases, and Google Scholar from December 01, 2019 to June 1, 2020. From an initial 547 publications and 75 reports, 25 studies provided specific information on COVID-19 patients living with HIV. Studies described 252 patients, 80.9% were male, mean age was 52.7 years, and 98% were on ART. Co-morbidities in addition to HIV and COVID-19 (multimorbidity) included hypertension (39.3%), obesity or hyperlipidemia (19.3%), chronic obstructive pulmonary disease (18.0%), and diabetes (17.2%). Two-thirds (66.5%) had mild to moderate symptoms, the most common being fever (74.0%) and cough (58.3%). Among patients who died, the majority (90.5%) were over 50 years old, male (85.7%), and had multimorbidity (64.3%). Our findings highlight the importance of identifying co-infections, addressing co-morbidities, and ensuring a secure supply of ART for PLHIV during the COVID-19 pandemic.

## Introduction

Coronavirus disease 2019 (COVID-19) was declared a pandemic on March 11^th^, 2020 (1). As of July 10^th^, 2020, 12,102,328 COVID-19 patients and 551,046 COVID-10-related deaths have been reported worldwide (2). The World Health Organization (WHO) and Center for Disease Control and Prevention (CDC) have issued health alerts and prevention guidelines for people at increased risk for severe health outcomes and death due to COVID-19 (3, 4). These guidelines are generally based on the outcomes and characteristics of patients affected early in the course of the COVID-19 pandemic. Emerging patterns point to elevated risk for older persons, people living in long-term care facilities, men, and racial/ethnic minorities who have long experienced disparities in health outcomes for many chronic diseases (5). Chronic disease co-morbidities, especially multimorbidity, appear to be driving factors for COVID-19 mortality. Warnings to take extra precautions include persons with asthma, chronic lung disease, diabetes, serious cardiovascular conditions, chronic kidney disease, obesity, chronic liver disease, and persons who are immunocompromised such as people living with HIV (PLHIV) (3–5).

The concern over increased risk for severe COVID-19 disease for PLHIV may be based on the assumption that PLHIV are more likely to be immunosuppressed. HIV infection is associated with abnormal humoral and T-cell–mediated immune responses, resulting in increased susceptibility to numerous opportunistic infections (6). Under this rationale, particular caution is warranted for PLHIV with low CD4 cell count, advanced disease, high viral load, and those not taking antiretroviral treatment (ART). As PLHIV are living longer with ART, many will also have the known chronic conditions (7) associated with severe COVID-19 disease. However, there have not been large observational studies specifically measuring symptoms, disease severity, complications, multimorbidity, and proportion of death in confirmed COVID-19-HIV co-infected patients. It is also not known if people with HIV who are clinically and virologically stable will experience any greater risk for COVID-19 complications than the population without HIV infection. At present, the available data mainly appear in case reports and case series of COVID-19-HIV co-infected patients.

Given the urgency of the COVID-19 pandemic and the rapidly changing information about the disease, a high degree of vigilance is needed on the course of infection among PLHIV. As there are 37.9 million PLHIV and 1.7 million new infections each year (8), patients of COVID-19-HIV co-infection are likely to increase. This systematic review was therefore undertaken to bring together the existing evidence on the earliest known case reports to provide a baseline for what is known and to alert providers around the world of any emerging patterns.

## Methods

Details of inclusion criteria and our analytical approach were conceptualized *a priori* and are documented in Open Science Framework (https://osf.io/zj2hu/).

### Literature search

Following the Systematic Reviews and Meta-Analyses (PRISMA) checklist (see supplementary file S1) and the Peer Review of Electronic Search Strategies (9) guideline (10, 11), we searched PubMed, Scopus, Web of Science, Embase, preprint databases (medRxiv, bioRxiv, Preprints), the references of publications found, and the “cited by” feature of Google Scholar from December 1, 2019 to June 1, 2020 for studies published in English. Search terms were combined using appropriate Boolean operators and included subject heading terms/keywords relevant to COVID-19 (e.g., SARS-CoV-2 OR Coronavirus Disease 2019 OR COVID-19 OR severe acute respiratory syndrome coronavirus 2 OR coronavirus infection) and HIV (e.g., HIV OR human immunodeficiency virus OR AIDS OR Acquired Immunodeficiency Syndrome). Please see supplementary file S2 for a sample search strategy.

### Inclusion criteria and study selection

Empirical studies including any study design (i.e., case report, case series, cross-sectional, case-control, cohort, and clinical trial) that reported individual- or aggregate-level data on COVID-19 among PLHIV were considered for this review. Studies that included a mixed sample of HIV-positive and HIV-negative COVID-19 patients were only considered if subgroup analyses for PLHIV were reported or could be extracted. Studies were excluded if they did not present original empirical data (e.g., editorials, commentaries, letters to editors, and reviews) or did not report any clinical data on patients with HIV and COVID-19 co-infection. The title, abstract, and full-text screening were completed in duplicate and independently by two reviewers. Duplicate records were excluded and disagreements over the inclusion of studies for data extraction were resolved through discussion or feedback from the senior author.

### Data extraction

Data extraction was completed in duplicate and discrepancies were resolved through discussion or feedback from the senior author. Data extracted included publication date, study type, location, sample size, participants’ age and sex, as well as HIV-related data such as CD4 count (cells/mm3), viral load (copies/ml), and antiretroviral therapy before COVID-19 diagnosis. COVID-19 related data including clinical symptoms (e.g., fever, cough, myalgia, dyspnea, headache, sore throat, fatigue, and gastrointestinal symptoms). Comorbidities other than HIV included atrial fibrillation, chronic kidney disease, congestive heart failure, chronic liver disease, cerebrovascular accident, cardiovascular disease, dyslipidemia, diabetes mellitus, hyperlipidemia, hypertension, obstructive sleep apnea, pulmonary embolism, obesity, and smoking. Severity of COVID-19 disease was classified as mild (i.e., non-pneumonia and mild pneumonia), severe (i.e., dyspnea, respiratory frequency□≥□30/min, blood oxygen saturation□≤□93%, and/or lung infiltrates□>□50% within 24–48 h), and critical (i.e., respiratory failure, septic shock, and/or multiple organ dysfunction or failure) (12). Admission to the intensive care unit (ICU) and recovery status (cured, died, still in hospital) were also collected.

### Quality assessment

The Joanna Briggs Institute critical appraisal tools were used to assess the methodological quality of the included papers (13). Selected studies were examined for inclusion criteria, sample size, description of study participants, setting, and the appropriateness of the statistical analysis. Methodological quality was independently assessed by two reviewers and disagreements were resolved through discussion. The tool was modified to provide a numeric score (14, 15). Quality assessments were done with different tools based on different study designs. Tools had nine items for case reports, ten items for case series, nine items for cross-sectional studies, and eleven items for cohort studies. Quality assessment tools and scores are presented in supplementary file S3.

### Statistical analysis

Descriptive analyses were used for reporting results. Continuous variables were summarized using mean and standard deviation (SD) with differences compared using a two-tailed student’s t-test. Categorical variables were summarized by frequency and percentage and differences were measured using the Fisher’s exact test. P-values less than 0.05 were considered as statistically significant. For combining data from studies that reported aggregated data with those reported individual data, aggregated data were weighted by the number of patients. The proportion of death among reported patients in studies included in the review was also measured. The denominator and nominator for this measure are based on people living with HIV whose COVID-19 was diagnosed and reported. We also conducted a subgroup analysis by sex.

## Results

The combined search strategy identified 622 potential publications on COVID-19 infection among PLHIV (Fig 1). After screening and removing duplicate studies (n = 222) and those not relevant to COVID-19-HIV co-infection (n = 343), the full text of 57 reports were sought to assess for eligibility. Of these, eight did not reported information about patients with co-infection; 23 were editorials, commentaries, or reviews; and the full-text for one abstract was unavailable. The search found 25 studies that met our inclusion criteria and were included in the systematic review (Table 1).

**Fig 1.**
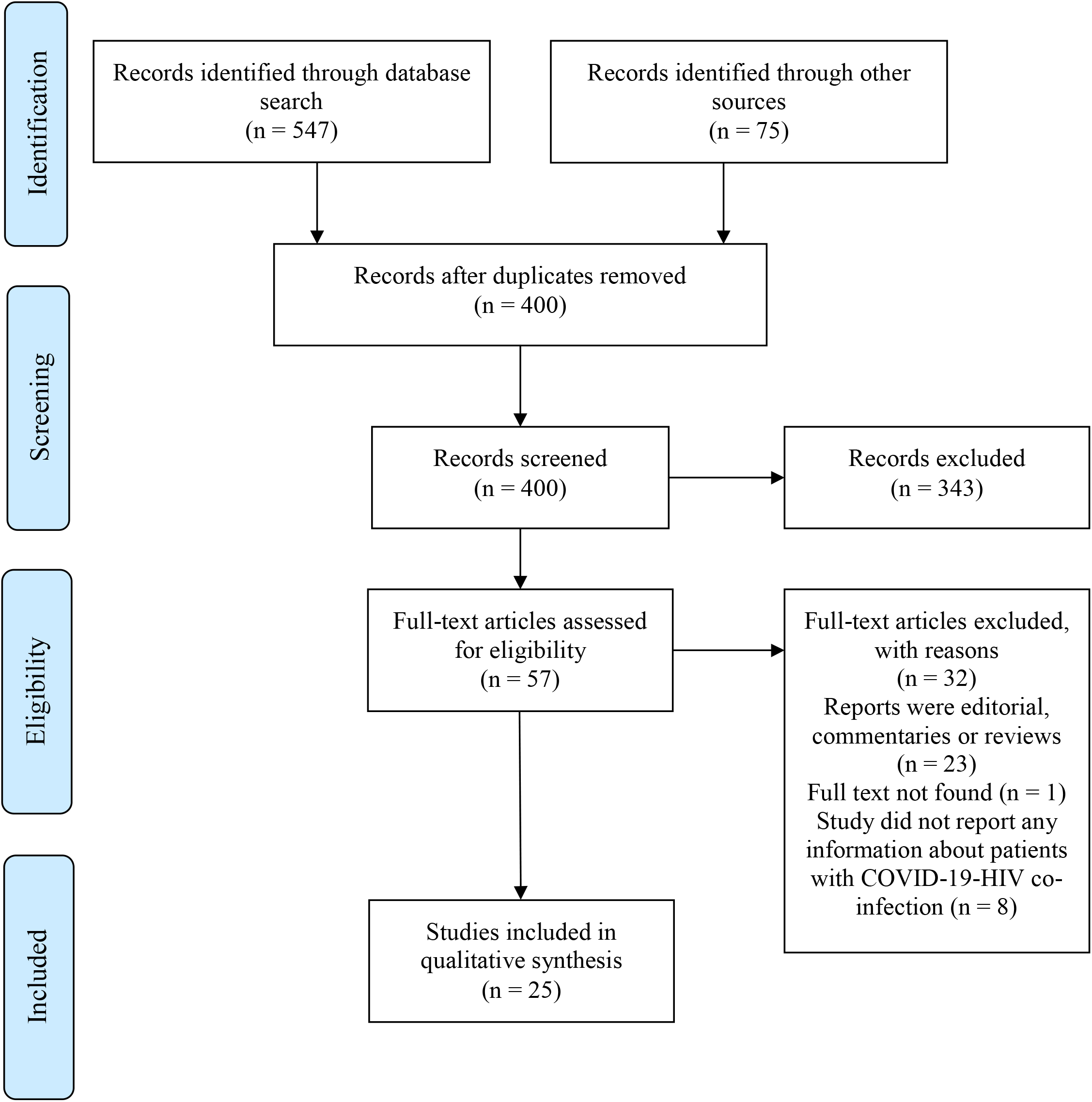
Flowchart of studies included in the systematic review of COVID-19-HIV co-infection

**Table 1.** Characteristic of studies included in the review of COVID-19-HIV co-infection.

| First author (ref) | Setting | Publication Date | Study Type | Data Type* | Sample Size | Case Definition | Quality Assessment |
| --- | --- | --- | --- | --- | --- | --- | --- |
| Zhu (22) | Wuhan, China | 12/03/20 | Case report | Individual | 1 | Confirmed | 5/8 |
| Guo (23) | Wuhan, China | 03/04/20 | Cross sectional | Aggregate | 8 | Confirmed | 7/9 |
| Zhao (24) | Shenzhen, China | 10/04/20 | Case report | Individual | 1 | Confirmed | 6/8 |
| Chen (25) | Guizhou, China | 15/04/20 | Case report | Individual | 1 | Confirmed | 6/8 |
| Su (26) | China | 17/04/20 | Case report | Individual | 1 | Confirmed | 6/8 |
| Schweitzer (27) | Italy | 18/04/20 | Case report | Individual | 1 | Confirmed | 4/8 |
| Blanco (28) | Barcelona, Spain | 19/04/20 | Case series | Individual | 5 | Confirmed | 8/10 |
| Riva (29) | Italy | 24/04/20 | Case series | Individual | 3 | Confirmed | 4/10 |
| Wang (30) | Wuhan, China | 27/04/20 | Case report | Individual | 1 | Confirmed | 6/8 |
| Altuntas Aydin (31) | Istanbul, Turkey | 30/04/20 | Case series | Individual | 4 | Confirmed | 7/10 |
| Haerter (32) | Germany | 01/05/20 | Case series | Individual | 33 | Confirmed | 8/10 |
| Karmen (33) | New York, USA | 12/05/20 | Retrospective cohort | Aggregate | 21 | Confirmed | 8/11 |
| Wu (34) | Wuhan, China | 13/05/20 | Case series | Individual | 2 | Confirmed | 4/10 |
| Gervasoni (35) | Italy | 15/05/20 | Cross sectional | Aggregate | 47 | Confirmed / Probable | 6/9 |
| Benkovic (36) | New York, USA | 20/05/20 | Case series | Individual | 4 | Confirmed | 4/10 |
| Haddad (37) | Wynnewood, USA | 20/05/20 | Case report | Individual | 1 | Confirmed | 7/8 |
| Baluku (38) | Uganda | 22/05/20 | Case report | Individual | 1 | Confirmed | 6/8 |
| Patel (39) | USA | 23/05/20 | Case report | Individual | 1 | Confirmed | 6/8 |
| Iordanou (40) | Cyprus | 25/05/20 | Case report | Individual | 1 | Confirmed | 7/8 |
| Kumar (41) | Chicago, USA | 27/05/20 | Case report | Individual | 1 | Confirmed | 7/8 |
| Childs (42) | UK | 28/05/20 | Case series | Aggregate | 18 | Confirmed | 4/10 |
| Suwanwongse (43) | New York, USA | 29/05/20 | Case series | Individual | 9 | Confirmed | 5/10 |
| Ridgway (44) | Chicago, USA | 30/05/20 | Case series | Individual | 5 | Confirmed | 7/10 |
| Shalev (45) | New York, USA | 31/05/20 | Case series | Aggregate | 31 | Confirmed | 8/10 |
| Vizcarra (20) | Madrid, Spain | 01/06/20 | Prospective cohort | Aggregate | 51 | Confirmed , probable | 9/11 |

\*Individual: reported information for each patient; Aggregate: reported summary of information for groups of patients

Of the 25 studies on COVID-19-HIV co-infection, 19 reported information on an individual-level, and six reported on an aggregate-level. Eleven studies were case reports (i.e., describing one patients), ten studies were case series including two to 33 patients. Two studies were cross sectional investigating COVID-19 status among PLHIV. Two cohort studies were found. Eight studies were from the US, seven from China, three from Italy, two from Spain, and the remaining five studies were from Turkey, Germany, the UK, Republic of Cyprus, and Uganda. Publication dates ranged from 12 March to 1 June, 2020. The Joanna Briggs Institute critical appraisal tools assessment scores ranged from 4 to 7 for case reports (out of 8 possible points), and 4 to 8 for the case series (out of 10 possible points), 6 to 7 for cross sectional (out of 9 possible points), and 8 to 9 for cohort studies (out of 11 possible points). As quality assessments were done with different tools based on different study designs, scores cannot not be directly compared.

Summed across all studies, 252 patients with COVID-19-HIV co-infection were described with varying completeness of demographic and clinical data (Table 2). The mean age of patients was 52.7 years. Most patients (204, 80.9%) were male; 46 (18.3%) were female, and two (0.8%) were transgender women. Of 251 patients in studies reporting ART status, only 2.0% were not taking ART. Low CD4 count (<200 cells/mm^3^) was reported for 23 of 176 patients (13.1%). The viral load was high (>1000 copies per ml) for two of 233 patients (0.9%) with viral load data. Multimorbidity included hypertension (96 of 244 patients reporting on co-morbidities, 39.3%), obesity or hyperlipidemia (as reported, 19.3%), chronic obstructive pulmonary disease (18.0%), and diabetes (17.2%). Smoking was reported in 53 patients (21.7%).

**Table 2.** Demographic and clinical characteristics of COVID-19 infection in people living with HIV included in the reviewed studies (N = 252 total patients)

| **Characteristics (n reported)\*** | **N (%)** |
| --- | --- |
| **Age (Mean ± SD; n = 244)** | 52.7 ± 8.6 |
| **Sex (n = 252)** |  |
| Male | 204 (80.9) |
| Female | 46 (18.3) |
| Transgender | 2 (0.8) |
| **On antiretroviral treatment (n = 251)** |  |
| Yes | 246 (98.0) |
| **CD4 count [cells/mm<sup>3</sup>] (n = 176)** |  |
| < 200 | 23 (13.1) |
| ≥ 200 | 153 (86.9) |
| **HIV viral load [copies per ml] (n = 233)** |  |
| ≤ 1000 | 231 (99.1) |
| > 1000 | 2 (0.9) |
| **Co-morbidities (in addition to COVID-19-HIV, n = 244)** |  |
| At least one morbidity | 145 (59.4) |
| Hypertension | 96 (39.3) |
| Obesity or hyperlipidemia | 47 (19.3) |
| Chronic obstructive pulmonary disease | 44 (18.0) |
| Diabetes | 42 (17.2) |
| Cardiovascular disease | 26 (10.7) |
| Renal insufficiency | 29 (11.9) |
| HBV/HCV | 18 (7.4) |
| Hypothyroidism | 1 (0.4) |
| Smoking | 53 (21.7) |
| **COVID-19 symptoms (n = 223)** |  |
| Fever | 165 (74.0) |
| Cough | 130 (58.3) |
| Dyspnea | 68 (30.5) |
| Headache | 44 (19.7) |
| Arthralgia/myalgia | 33 (14.8) |
| Sore throat | 18 (8.1) |
| Gastrointestinal symptoms | 29 (13.0) |
| **Severity (n = 212)** |  |
| Mild or moderate | 141 (66.5) |
| Severe | 46 (21.7) |
| Critical | 25 (11.8) |
| **Hospitalized (n = 244)** |  |
| Yes | 158 (64.7) |
| **Intensive care unit admission (n = 244)** |  |
| Yes | 41 (16.8) |
| **Death (n = 252)** |  |
| Yes | 36 (14.3) |

\*Continuous variables were summarized using mean and standard deviation and categorical variables were summarized using frequency and percentage. Data are n (%) unless otherwise stated.

Table 2 also presents reported symptoms and severity of COVID-19 disease patients with HIV infection. The most common symptoms were fever (165 of 223, 74.0%), cough (130 of 223, 58.3%), and dyspnea (68 of 223, 30.5%). Less common were headache (44 of 223, 19.7%), arthralgia/myalgia (33 of 223, 14.8%), and sore throat (18 of 223, 8.1%). Any gastrointestinal symptoms was reported by 13.0%. COVID-19 was reported as mild to moderate in 141 of 212 (66.5%), severe in 46 patients (21.7%), and critical in 25 patients (11.8%). The majority of patients (158 of 244, 64.7%) were hospitalized; 16.8% were admitted to the intensive care unit.

Of all 252 reported cases of COVID-19-HIV co-infection, 36 (14.3%) were reported as having died. Supplementary file S4 presents further information on patients where available. Information on the sex of the deceased were available for 14 of 36 patients, with 85.7% being male. Information on the age of the deceased was available for 21 of 36 patients, with 38.1% of deaths occurring in patients over 65 years of age and older, 52.4% in patients age 50 to 65 years of age, and 9.5% in patients younger than 50 years. Information about multimorbidity, was available for 14 of 36 the deceased patients, with 64.3% reporting multimorbidity. Additional information on ART regimen, CD4 count, viral load, and indicators of severity of disease can be found in supplementary file S4.

Based on the available data, no significant differences between male and female patients were observed with regards to age, clinical characteristics, severity of COVID-19, and health outcomes of COVID-19-HIV co-infected patients (Table 3).

**Table 3.**
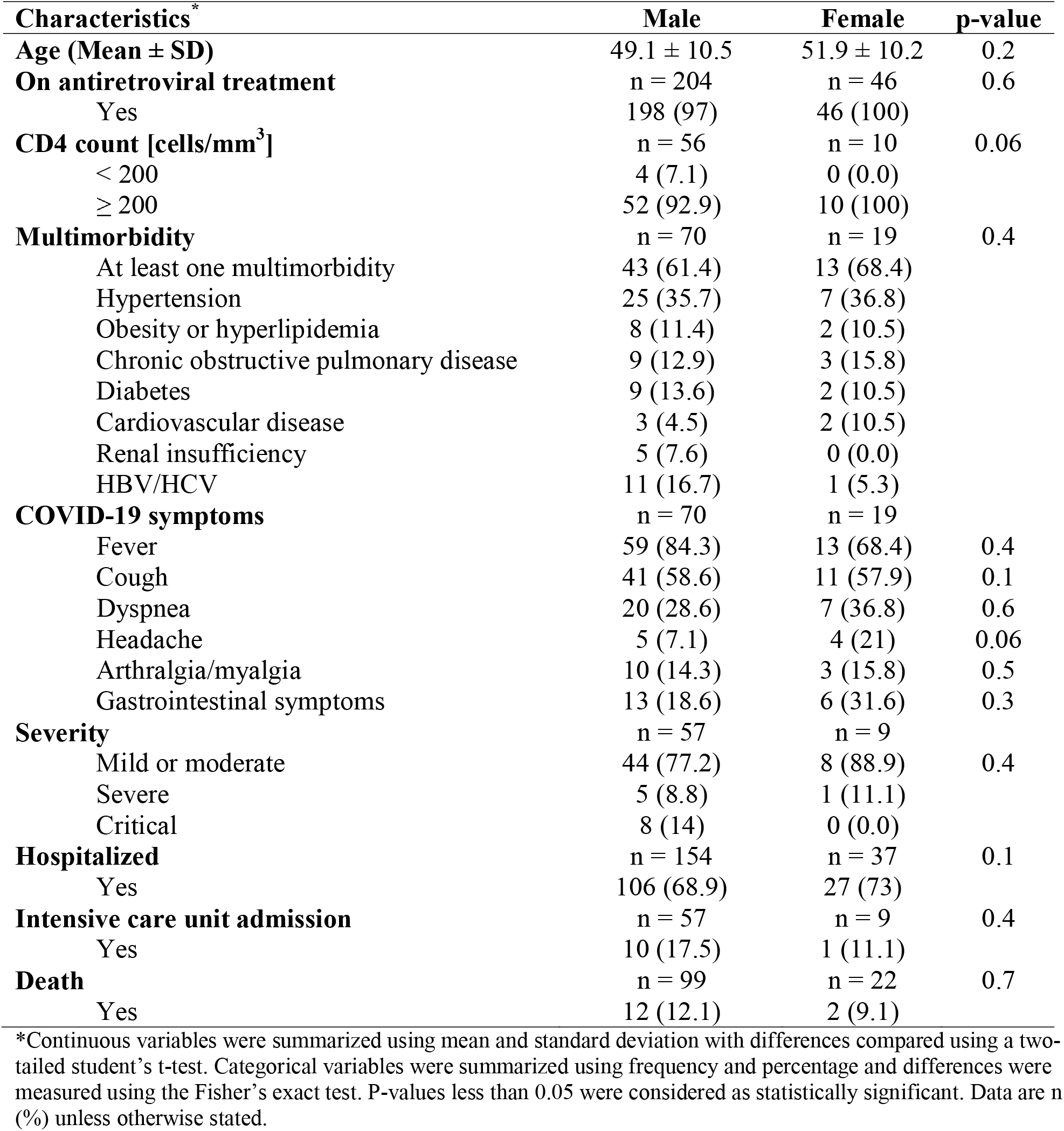
Demographic and clinical characteristics of COVID-19 in people living with HIV included in the reviewed studies, stratified by sex.

| **Characteristics\*** | **Male** | **Female** | **p-value** |
| --- | --- | --- | --- |
| **Age (Mean ± SD)** | 49.1 ± 10.5 | 51.9 ± 10.2 | 0.2 |
| **On antiretroviral treatment** | n = 204 | n = 46 | 0.6 |
| Yes | 198 (97) | 46 (100) |  |
| **CD4 count [cells/mm<sup>3</sup>]** | n = 56 | n = 10 | 0.06 |
| < 200 | 4 (7.1) | 0 (0.0) |  |
| ≥ 200 | 52 (92.9) | 10 (100) | |
| **Multimorbidity** | n = 70 | n = 19 | 0.4 |
| At least one multimorbidity | 43 (61.4) | 13 (68.4) |  |
| Hypertension | 25 (35.7) | 7 (36.8) |  |
| Obesity or hyperlipidemia | 8 (11.4) | 2 (10.5) |  |
| Chronic obstructive pulmonary disease | 9 (12.9) | 3 (15.8) |  |
| Diabetes | 9 (13.6) | 2 (10.5) |  |
| Cardiovascular disease | 3 (4.5) | 2 (10.5) |  |
| Renal insufficiency | 5 (7.6) | 0 (0.0) |  |
| HBV/HCV | 11 (16.7) | 1 (5.3) |  |
| **COVID-19 symptoms** | n = 70 | n = 19 |  |
| Fever | 59 (84.3) | 13 (68.4) | 0.4 |
| Cough | 41 (58.6) | 11 (57.9) | 0.1 |
| Dyspnea | 20 (28.6) | 7 (36.8) | 0.6 |
| Headache | 5 (7.1) | 4 (21) | 0.06 |
| Arthralgia/myalgia | 10 (14.3) | 3 (15.8) | 0.5 |
| Gastrointestinal symptoms | 13 (18.6) | 6 (31.6) | 0.3 |
| **Severity** | n = 57 | n = 9 |  |
| Mild or moderate | 44 (77.2) | 8 (88.9) | 0.4 |
| Severe | 5 (8.8) | 1 (11.1) |  |
| Critical | 8 (14) | 0 (0.0) |  |
| **Hospitalized** | n = 154 | n = 37 | 0.1 |
| Yes | 106 (68.9) | 27 (73) |  |
| **Intensive care unit admission** | n = 57 | n = 9 | 0.4 |
| Yes | 10 (17.5) | 1 (11.1) |  |
| **Death** | n = 99 | n = 22 | 0.7 |
| Yes | 12 (12.1) | 2 (9.1) |  |

\*Continuous variables were summarized using mean and standard deviation with differences compared using a two-tailed student's t-test. Categorical variables were summarized using frequency and percentage and differences were measured using the Fisher's exact test. P-values less than 0.05 were considered as statistically significant. Data are n (%) unless otherwise stated.

## Discussion

In this systematic review, we summarized available data from 252 patients co-infected with COVID-19 and HIV. The majority of patients with co-infection of HIV and COVID-19 were male. Also in line with HIV-negative persons (16–19), data point to higher morbidity and mortality among male patients, and higher mortality when multimorbidity is present. The main clinical symptoms of COVID-19 in PLHIV were cough and fever, and comparable to HIV-negative people, the majority had mild COVID-19 disease (16–19).

Nonetheless, the proportion of reported PLHIV with COVID-19 appear to have higher multimorbidity, severity of disease, and potentially higher proportion of death. Multimorbidity was reported in nearly two-thirds of co-infected patients. The most common co-morbidities among patients with HIV and COVID-19 were hypertension, obesity or hyperlipidemia, COPD, and diabetes. Results of a cohort study showed that multimorbidity (mostly hypertension and diabetes) was more prevalent in COVID-19-HIV co-infected patients than PLHIV without COVID-19 (20). When data were available, the proportion of death among reported COVID-19-HIV co-infected patients appears high (14.3%) in our pooled estimate. The result might be confounded by other factors. For example, PLHIV may be at increased risk of mortality or severe illness with COVID-19 based on their age and other medical conditions. Among those who died for whom individual data were recorded, multimorbidity and older age were common. Current clinical data suggest the main mortality risk factors are linked to older age and multimorbidity not particular to HIV, including cardiovascular disease, diabetes, chronic respiratory disease, and hypertension (21). Another possible explanation is bias due to the preponderance of publications on hospitalized patients with more severe disease.

The observational designs of the available studies and lack of appropriate control do not permit concluding on whether ART can prevent acquisition of COVID-19 or reduce morbidity and mortality from COVID-19. The search for anti-viral agents with activity to treat and prevent COVID-19 is an area of active research. For example, a clinical trial in 199 patients showed that ritonavir-boosted lopinavir did not have a benefit over standard care for COVID-19 (15). A survey in PLHIV in China showed that nucleoside reverse transcriptase inhibitors (NRTI) plus non-nucleoside reverse transfer inhibitors (NNRTI) did not prevent COVID-19 infection (16). A cohort study showed that there are no differences in previous use of NNRTI, INSTI, or protease inhibitors in individuals with and without COVID-19 diagnosis (17). Findings of lower morbidity or mortality among persons on ART compared to those not on ART would also need to consider higher CD4 count and better immune status.

We acknowledge additional limitations of our study. First, while a large proportion of COVID-19 patients are asymptomatic or have mild symptoms, all of the patients in this review were symptomatic, and most were admitted to a hospital. This study therefore refers only to patients with confirmed diagnosis of COVID-19 and may over-represent those with severe COVID-19 disease. Second, the lack of appropriate comparison groups, particularly for the case reports and case series, is a limitation to identifying factors associated with COVID-19-HIV co-infection. Third, many of the studies included in our review had small sample size. Fourth, without a population- or probability-based survey of PLHIV, the true prevalence of COVID-19 and disease manifestations among PLHIV remain unknown. To the best of our knowledge, no study has yet measured COVID-19-HIV co-infection in a wider or representative cross-section of the population. Fifth, since most of the patients included in the studies reviewed were immunologically and virologically stable, we could not conduct subgroup analyses based on low CD4 cell count or high viral load to directly measure the association of HIV-related immunosuppression and severity of COVID-19 disease. Such subgroup analyses should be investigated in future studies. Finally, in the context of urgent care for large numbers of patients, HIV may not be divulged, asked for, or recorded and therefore under-assessed among COVID-19 patients.

## Conclusions

The data available to date indicate that PLHIV can be infected with COVID-19 and are largely affected by similar features of disease risk and progression as HIV-uninfected patients. The presence of multimorbidity and older age appear to be the important factors for severe morbidity and mortality with COVID-19-HIV co-infection. Results suggest that healthcare providers need to address multimorbidity among PLHIV, ensure their ART supply is secure, and continue to consider PLHIV as a population for whom precautions are needed to prevent the COVID-19. Given the number of PLHIV worldwide, there are likely many more patients with COVID-19-HIV co-infection than suggested by the scant literature to date. Clinicians and researchers could help fill the data gap by being vigilant to patients who may be co-infected with SARS-CoV-2 and HIV.

## Data Availability

All data is presented are the study

## Author contributions

All authors contributed to the study conception and design. Material preparation, data collection, and analysis were performed by HS and HM. The first draft of the manuscript was written by HM. WM and MK assisted with conceptualization, interpretation, composition, editing, and responding to the reviewers’ comments. All authors commented on previous versions of the manuscript. All authors read and approved the final manuscript.

## Acknowledgements

The authors did not receive any funding for this study. MK is a member of Pierre Elliott Trudeau Foundation’s COVID-19 impact committee and is supported by the Vanier Canada Graduate Scholarship and the Pierre Elliott Trudeau Foundation Doctoral Scholarship.

## Conflict of interests

The authors declare that they have no competing interests.

## Notes

### Competing Interest Statement

The authors have declared no competing interest.

### Clinical Trial

https://osf.io/zj2hu/

### Clinical Protocols

https://osf.io/zj2hu/

### Author Declarations

NA; This is a systematic review

## References

1. Zhu N, Zhang D, Wang W, Li X, Yang B, Song J, et al. A novel coronavirus from patients with pneumonia in China, 2019. New England Journal of Medicine. 2020.

2. WHO. WHO Coronavirus Disease (COVID-19) Dashboard [updated 10 July 2020. Available from: https://covid19.who.int/.

3. CDC. Coronavirus Disease 2019 (COVID-19) [updated 2020/6/2. Available from: https://www.cdc.gov/coronavirus/2019-ncov/cases-updates/cases-in-us.html.

4. WHO. Coronavirus disease (COVID-19) pandemic [updated 2020/6/3. Available from: https://www.who.int/emergencies/diseases/novel-coronavirus-2019.

5. Khalili M, Karamouzian M, Nasiri N, Javadi S, Mirzazadeh A, Sharifi H. Epidemiological characteristics of COVID-19: a systematic review and meta-analysis. Epidemiology and Infection. 2020;148:e130.

6. Chang CC, Crane M, Zhou J, Mina M, Post JJ, Cameron BA, et al. HIV and co□infections. Immunological reviews. 2013;254(1):114–42.

7. Garrido-Hernansaiz H, Heylen E, Bharat S, Ramakrishna J, Ekstrand ML. Stigmas, symptom severity and perceived social support predict quality of life for PLHIV in urban Indian context. Health and quality of life outcomes. 2016;14(1):152.

8. Mahy M, Marsh K, Sabin K, Wanyeki I, Daher J, Ghys PD. HIV estimates through 2018: data for decision-making. LWW; 2019.

9. Pressman P, Clemens R, Sahu S, Hayes AW. A Review of Methanol Poisoning: A Crisis Beyond Ocular Toxicology. Cutaneous and ocular toxicology. 2020:1–19.

10. Moher D, Liberati A, Tetzlaff J, Altman DG, Group P. Preferred reporting items for systematic reviews and meta-analyses: the PRISMA statement. PLoS med. 2009;6(7):e1000097.

11. McGowan J, Sampson M, Salzwedel DM, Cogo E, Foerster V, Lefebvre C. PRESS peer review of electronic search strategies: 2015 guideline statement. Journal of clinical epidemiology. 2016;75:40–6.

12. Wu Z, McGoogan JM. Characteristics of and Important Lessons From the Coronavirus Disease 2019 (COVID-19) Outbreak in China: Summary of a Report of 72□314 Cases From the Chinese Center for Disease Control and Prevention. JAMA. 2020;323(13):1239–42.

13. Joanna Briggs Institute. The Joanna Briggs Institute Critical Appraisal tools for use in J BI Systematic Reviews Checklist for Prevalence Studies.2017. Available at : http://joannabriggs.org/research/critical-appraisal-tools.html.

14. Karamouzian M, Nasirian M, Hoseini SG, Mirzazadeh A. HIV and Other Sexually Transmitted Infections Among Female Sex Workers in Iran: A Systematic Review and Meta-Analysis. Archives of sexual behavior. 2019:1–15.

15. Xu Y, Chen X, Wang K. Global prevalence of hypertension among people living with HIV: a systematic review and meta-analysis. Journal of the American Society of Hypertension. 2017;11(8):530–40.

16. Livingston E, Bucher K. Coronavirus Disease 2019 (COVID-19) in Italy. JAMA. 2020;323(14):1335-.

17. Chen N, Zhou M, Dong X, Qu J, Gong F, Han Y, et al. Epidemiological and clinical characteristics of 99 cases of 2019 novel coronavirus pneumonia in Wuhan, China: a descriptive study. The Lancet. 2020;395(10223):507–13.

18. Guan W-j, Ni Z-y, Hu Y, Liang W-h, Ou C-q, He J-x, et al. Clinical characteristics of coronavirus disease 2019 in China. New England journal of medicine. 2020;382(18):1708–20.

19. Richardson S, Hirsch JS, Narasimhan M, Crawford JM, McGinn T, Davidson KW, et al. Presenting Characteristics, Comorbidities, and Outcomes Among 5700 Patients Hospitalized With COVID-19 in the New York City Area. JAMA. 2020.

20. Vizcarra P, Pérez-Elías MJ, Quereda C, Moreno A, Vivancos MJ, Dronda F, et al. Description of COVID-19 in HIV-infected individuals: a single-centre, prospective cohort. The lancet HIV. 2020.

21. WHO. Clinical management of severe acute respiratory infection when novel coronavirus (nCoV) infection is suspected: interim guidance, 25 January 2020. World Health Organization; 2020.

22. Zhu F, Cao Y, Xu S, Zhou M. Co□infection of SARS□CoV□2 and HIV in a patient in Wuhan city, China. Journal of Medical Virology. 2020.

23. Guo W, Ming F, Dong Y, Zhang Q, Zhang X, Mo P, et al. A survey for COVID-19 among HIV/AIDS patients in two Districts of Wuhan, China. AIDS Patients in Two Districts of Wuhan, China (3/4/2020). 2020.

24. Zhao J, Liao X, Wang H, Wei L, Xing M, Liu L, et al. Early virus clearance and delayed antibody response in a case of COVID-19 with a history of co-infection with HIV-1 and HCV. Clinical infectious diseases : an official publication of the Infectious Diseases Society of America. 2020.

25. Chen J, Cheng X, Wang R, Zeng X. Computed Tomography Imaging of an HIV-infected Patient with Coronavirus Disease 2019 (COVID-19). J Med Virol. 2020.

26. Su J, Shen X, Ni Q, Zhao H, Cai J, Zhu B, et al. Infection of severe acute respiratory syndrome coronavirus 2 in a patient with acquired immunodeficiency syndrome. AIDS. 2020.

27. Schweitzer W, Ruder T, Baumeister R, Bolliger S, Thali M, Meixner E, et al. Implications for forensic death investigations from first Swiss post-mortem CT in a case of non-hospital treatment with COVID-19. For Imag. 2020;21.

28. Blanco JL, Ambrosioni J, Garcia F, Martinez E, Soriano A, Mallolas J, et al. COVID-19 in patients with HIV: clinical case series. The lancet HIV. 2020.

29. Riva A, Conti F, Bernacchia D, Pezzati L, Sollima S, Merli S, et al. Darunavir does not prevent SARS-CoV-2 infection in HIV patients. Pharmacological research. 2020;157:104826.

30. Wang M, Luo L, Bu H, Xia H. Case Report: One Case of Coronavirus Desease 2019 (COVID-19) in Patient Co-nfected by HIV With a Low CD4+ T Cell Count. International Journal of Infectious Diseases. 2020.

31. Altuntas Aydin O, Kumbasar Karaosmanoglu H, Kart Yasar K. HIV/SARS-CoV-2 co-infected patients in Istanbul, Turkey. J Med Virol. 2020.

32. Haerter G, Spinner CD, Roider J, Bickel M, Krznaric I, Grunwald S, et al. COVID-19 in people living with human immunodeficiency virus: A case series of 33 patients. medRxiv. 2020.

33. Karmen-Tuohy S, Carlucci PM, Zervou FN, Zacharioudakis IM, Rebick G, Klein E, et al. Outcomes among HIV-positive patients hospitalized with COVID-19. Journal of acquired immune deficiency syndromes (1999). 2020.

34. Wu Q, Chen T, Zhang H. Recovery from COVID□19 in two patients with coexisted HIV infection. Journal of Medical Virology. 2020.

35. Gervasoni C, Meraviglia P, Riva A, Giacomelli A, Oreni L, Minisci D, et al. Clinical features and outcomes of HIV patients with coronavirus disease 2019. Clinical infectious diseases : an official publication of the Infectious Diseases Society of America. 2020.

36. Benkovic S, Kim M, Sin E. 4 Cases: HIV and SARS-CoV-2 Co-infection in patients from Long Island, New York. J Med Virol. 2020.

37. Haddad S, Tayyar R, Risch L, Churchill G, Fares E, Choe M, et al. Encephalopathy and seizure activity in a COVID-19 well controlled HIV patient. IDCases. 2020:e00814.

38. Baluku JB, Mwebaza S, Ingabire G, Nsereko C, Muwanga M. HIV and SARS-CoV-2 co-infection: A case report from Uganda. J Med Virol. 2020.

39. Patel RH, Pella PM. COVID-19 in a patient with HIV infection. J Med Virol. 2020.

40. Iordanou S, Koukios D, Matsentidou CT, Markoulaki D, Raftopoulos V. Severe SARS-CoV-2 pneumonia in a 58-year-old patient with HIV: a clinical case report from the Republic of Cyprus. J Med Virol. 2020.

41. Kumar RN, Tanna SD, Shetty AA, Stosor V. COVID-19 in an HIV-positive Kidney Transplant Recipient. Transplant infectious disease : an official journal of the Transplantation Society. 2020:e13338.

42. Childs K, Post FA, Norcross C, Ottaway Z, Hamlyn E, Quinn K, et al. Hospitalized patients with COVID-19 and HIV: a case series. Clinical infectious diseases : an official publication of the Infectious Diseases Society of America. 2020.

43. Suwanwongse K, Shabarek N. Clinical features and outcome of HIV/SARS-CoV-2 co-infected patients in the Bronx, New York City. J Med Virol. 2020.

44. Ridgway JP, Farley B, Benoit JL, Frohne C, Hazra A, Pettit N, et al. A Case Series of Five People Living with HIV Hospitalized with COVID-19 in Chicago, Illinois. AIDS patient care and STDs. 2020.

45. Shalev N, Scherer M, LaSota ED, Antoniou P, Yin MT, Zucker J, et al. Clinical characteristics and outcomes in people living with HIV hospitalized for COVID-19. Clinical infectious diseases : an official publication of the Infectious Diseases Society of America. 2020.

